# Reduced heart rate variability is associated with vulnerability to depression

**DOI:** 10.1101/2020.09.22.20199356

**Authors:** Carola Dell’Acqua, Elisa Dal Bò, Simone Messerotti Benvenuti, Daniela Palomba

## Abstract

**Background:** Heart rate variability (HRV) mirrors cardiac autonomic modulation, an index of well-being. Reduced HRV has been reported in depression, but few studies investigated HRV in individuals at-risk of or remitted from a full-blown depressive episode. The present study aimed at examining whether reduced HRV could be a potential indicator of vulnerability to depression.

**Methods:** Self-reported psychological measures and three-minute resting-state ECG were collected in two at-risk populations [group with dysphoria (*n* = 27), group with past depression (*n* = 16)] and in a control group (*n* = 25). Time- and frequency-domain HRV parameters were computed. Analysis of covariance was conducted to detect between-groups differences for each measure.

**Results:** Standard Deviation of Normal to Normal intervals (SDNN) and High Frequency (HF) power of HRV were found to be reduced both in individuals with dysphoria and in those with past depression as compared with controls. Whereas psychological measures did not significantly differ among individuals with past depression and controls, HRV was capable of discriminating between the two groups.

**Limitations:** Past depression was assessed retrospectively with self-reported information. The inclusion of a group with depression would provide an overview about HRV during the illness course.

**Conclusions:** The findings suggest that reduced HRV is likely to be implicated in the risk of developing full-blown depression, rather than being a mere correlate of current depressive state. The results suggest that HRV may improve clinicians’ ability to early identify people at risk for depression who can benefit from targeted prevention by psychiatric and psychological interventions.

## 1. Introduction

Major Depressive Disorder (MDD) is outlined as a mood disorder associated with a persistent state of sadness and/or loss of interest that affects thoughts and behavior, including physical and cognitive difficulties (Kessler et al., 2003). Considering the high prevalence and extensive burden of depression, many studies have been conducted with the aim to increase the understanding of symptoms, maintenance and possible interventions. Yet, so far, research has focused their investigations mainly on the acute phases of depressive symptoms, even though MDD is fundamentally known to be a chronic condition due to elevated episodes of relapse and recurrence (Richards, 2011). Indeed, research showed that the recurrence of depression is estimated to be about 35% after 15 years in the general population and about 60% after five years in those treated by mental health professionals (Hardeveld et al., 2010). Accordingly, the European Pact for Mental Health and Well-Being has highlighted as key priorities advancing research on the prevention and early recognition of depression (Henderson et al., 2004; Wahlbeck and Mäkinen, 2008). In this context, the need to identify vulnerability factors that contribute to depression onset or subclinical depression persistence becomes central. In the present work, two at-risk categories are taken into account: individuals who have suffered but are currently remitted from depressive symptoms (Hardeveld et al., 2010) and individuals with dysphoria (Meeks et al., 2011). Dysphoria, or subclinical depression, is a condition characterized by elevated depressive symptoms that do not meet the criteria for a formal diagnosis of MDD or dysthymia (Frewen and Dozois, 2005; Haaga and Solomon, 1993; Kendall et al., 1987) and is acknowledged to increase significantly the risk of developing a full-blown major depression or dysthymia (Horwath et al., 1992). Although the classic nosology of mental disorders has highly underestimated the importance of subthreshold conditions, it was demonstrated that dysphoria is characterized by significant difficulties in functioning that result in having a negative impact on the quality of life (Rodríguez et al., 2012).

Vulnerability to depressive occurrence and recurrence is almost certainly determined by a complex interaction of biological, psychological and environmental factors (Burcusa and Iacono, 2007). Several clinical predictors of depression occurrence and recurrence have been recognised, including age of illness onset, long-lasting subclinical symptoms, dysfunctional cognitive biases and emotion regulation strategies (Burcusa and Iacono, 2007; Miranda and Pearson 1988; Teasdale, 1988; ten Doesschate et al., 2010). Despite biological factors of depression have been broadly explored in acute phases of depression, their role in vulnerability to depression has not been properly investigated. This approach is misleading because any alterations during an acute episode could embody the clinical state itself and its treatment rather than the actual mechanisms involved in the pathophysiology (Bhagwagar and Cowen, 2007). Therefore, the need to focus on individuals who are at risk and more vulnerable to the development or persistence of a full-blown depressive episode becomes crucial in shedding light on psychobiological factors contributing to MDD onset.

Recently, a comprehensive approach has been employed in order to understand the complexity of the interacting systems that constitute the human body. The autonomic nervous system (ANS) balance seems to be a key component in the regulation of most of the physiological systems and its central control has been located in several portions of the brain (Ernst, 2017), in particular in the hypothalamus, the limbic cortex, the amygdala and the prefrontal cortex, making the Central Autonomic Network (CAN; Thayer et al., 2012). Within this framework, repeated findings reporting the high occurrence of autonomic cardiac alterations among individuals with depression have underlined the potential role of the ANS regulation in the pathophysiological mechanisms of depression (e.g., Hartmann et al., 2019). Several studies in this field support the notion that depression is characterized by an altered autonomic activity, with an increased sympathetic and/or a decreased parasympathetic nervous system activity (Gentili et al., 2017; Koch et al., 2019; Patron et al., 2012; Siever and Davis, 1985; Udupa et al., 2007).

Resting heart rate variability (HRV) is a commonly used measure to assess cardiac autonomic balance, as it reflects the balance between the two ANS branches on the heart (Task Force, 1996). Considering that the heart is under tonic inhibitory control via the vagus nerve and predominantly dominated by vagal influences, low HRV reflects reduced cardiac vagal control (Levy, 1990; Task Force, 1996; Thayer and Lane, 2000). A compelling body of research found that depression is characterized by lower time- and frequency-domain HRV parameters (for a meta-analysis see Kemp et al., 2010), and these measures have been used to identify MDD patients or healthy subjects (Hartmann et al., 2019; Jangpangi et al., 2016; Nahshoni et al., 2004). Also, the central role of the ANS in depression is highlighted by the polyvagal theory (Porges, 1995), which might represent an index of depression that has been poorly considered so far (Porges, 1995). Furthermore, considering that HRV is modulated by the CAN (Thayer et al., 2012), a reduction in HRV parameters could, in turn, imply dysfunctions in these brain pathways that persist after remission from depressive symptoms, or that could even be present before depression onset (Bassett et al., 2016).

Despite the evidence supporting the presence of reduced HRV during acute episodes of MDD (Koch et al., 2019; Udupa et al., 2007), studies addressing subclinical depression and remitted depression are lacking. To the best of our knowledge, only few studies have examined the reduction of HRV parameters in individuals with past depressive symptoms or dysphoria (Basset et al., 2016; Chang et al., 2013; Messerotti Benvenuti et al., 2015). For instance, a significant reduction in most of HRV parameters in individuals with past depression, despite the current absence of depressive symptoms, was reported (Basset et al., 2016). On the other hand, a study that employed a large sample of individuals with past depression found that decreased cardiac vagal control is only present in a limited subgroup of individuals with a history of suicidal ideation (Chang et al., 2013). Moreover, even though dysphoria-related HRV reductions have never been directly reported, a recent study found that somatic, but not cognitive-affective symptoms were inversely associated with HRV in a group of participants with dysphoria as compared to a control group (Messerotti Benvenuti et al., 2015). Additionally, a longitudinal study on a non-clinical sample reported that the presence of depressive symptoms was correlated with low vagally mediated HRV on a time frame of almost three years, suggesting that low vagal tone may be prospectively implicated in the generation of depression (Carnevali et al., 2018).

The aim of the present study was to investigate HRV parameters in individuals that are vulnerable to the onset of a full-blown depressive episode and, in turn, to understand whether reduced HRV could be associated with a higher risk for the development of clinical depression. To this end, two groups were enrolled in the present study: (1) individuals with dysphoria, a condition characterized by depressive symptoms without meeting the diagnostic criteria for MDD, and (2) individuals with unmedicated past depressive symptoms but currently free from clinical symptoms. Both groups represent samples of individuals that are currently free from clinical depression, still being more vulnerable to the development of a full-blown depressive episode than the general population (Hardeveld et al., 2010; Laborde-Lahoz et al., 2015; Meeks et al., 2011). It was hypothesized that individuals with dysphoria and individuals with past depressive symptoms would show reduced HRV parameters (time- and-frequency domain) compared to healthy controls.

## 2. Methods

### 2.1. Participants

A total of 68 university students voluntarily took part in the present study. The enrolled sample was medically healthy and free from psychotropic medication, as assessed with an ad-hoc anamnestic interview. The module A of the Structured Clinical Interview for DSM-5 (SCID 5-CV; First et al., 2016; Fossati and Borroni, 2017) was administered in order to assess current and past depressive symptoms and to exclude individuals with current major depression, persistent depressive disorder or bipolar disorder. Also, participants completed the Beck Depression Inventory-II (BDI-II; Beck et al., 1996; Ghisi et al., 2006) in order to assess the severity of depressive symptoms. Participants with a BDI-II score equal to or greater than 12 and at least two current depressive symptoms lasting minimum two weeks without meeting the diagnostic criteria for major depression, dysthymia or bipolar disorder, were assigned to the group with dysphoria (*n* = 27). Participants who met the criteria for past major depression episode, who had been in remission for minimum six months, with a BDI-II score lower than 8 and no current depressive symptoms, were included in the group with past depressive symptoms (PDS; *n* = 16). According to SCID-5-CV, past major depression episodes are characterized by at least two weeks of depressive symptoms, presenting at least either sad mood or anhedonia, that caused significant modifications in day-to-day functioning. Also, SCID-5-CV was useful to confirm full recovery from the depressive episode. A substantial number of studies have used both the BDI-II and the SCID-5-CV to classify past depressive episodes (e.g., Fritzsche et al., 2010; Stewart et al., 2011; Zvielli et al., 2016) and dysphoria (e.g., Balderas et al., 2019; Del Palacio-Gonzalez et al., 2017; Messerotti Benvenuti et al., 2017; Messerotti Benvenuti et al. 2019; Messerotti Benvenuti et al., 2020; Yang et al., 2020). In order to ensure separation between groups with and without dysphoria, participants with a BDI-II score lower than 8 and no history of depression or current depressive symptoms were included in the control group (*n* = 25). Participants were compensated 13 € for their participation. The present study was conducted with the adequate understanding and written consent of the participants in accordance with the Declaration of Helsinki and was approved by the local Ethics Committee, University of Padua (prot. no. 3612).

### 2.2. Psychological evaluation

Participants were administered the mood episode module (module A) of the SCID-5-CV to assess the presence of current and past depressive symptoms. The SCID-5-CV is a reliable tool to assess both subclinical symptoms and previous clinical symptoms in patients remitted from depression (internal consistency: Cronbach’s alpha = .91; Shankman et al., 2018). The BDI-II is a reliable and valid self-report questionnaire that was used to assess the severity of current depressive symptoms. Specifically, the BDI-II is a valid and reliable self-report questionnaire that evaluates the severity of depressive symptoms in the past two weeks and is composed of 21 items (internal consistency: Cronbach’s alpha = .87). It has a four-point Likert scale and scores range from 0 to 63, with the higher scores indicating greater depressive symptoms. In the Italian version, a score of 12 has been reported as the optimal cut-off score to discriminate individuals with and without depressive symptoms (Ghisi et al., 2006). Then, two questionnaires were administered in order to assess multiple aspects of participants’ affective states: The Beck Anxiety Inventory (BAI, Beck et al., 1988; Sica et al., 2006; internal consistency: Cronbach’s alpha = .89) was used to assess anxiety and the Emotion Regulation Questionnaire (ERQ, Balzarotti et al., 2010; Gross and John, 2003) was used to evaluate emotion regulation strategies of cognitive reappraisal (i.e., ERQ-Reappraisal, internal consistency: Cronbach’s alpha = .84) and suppression of emotion-expressive behavior (i.e., ERQ-Suppression, internal consistency: Cronbach’s alpha = .72).

### 2.3. Procedure

Prior to the experimental session, participants were requested to avoid alcohol consumption the day before and to avoid caffeine and nicotine the same day of the appointment. Upon arrival at the laboratory, after reading and signing the informed consent, participants were first administered a demographic interview and the mood episode module (module A) of the SCID-5-CV by a trained clinical psychologist. Then, participants were seated in a dimly lit, sound-attenuated room. After the sensors were attached, ECG was recorded at rest over a three-minute period for each participant. Ultra-short-term recordings (<5 minutes) have been documented to be a reliable method to measure HRV (Baek et al., 2015; Shaffer et al., 2016). Subsequently, participants completed the battery of the self-reported psychological measures. Following completion of the self-reported questionnaires, the participants were fully debriefed. The entire procedure lasted approximately 60 min.

### 2.4. Physiological recording

Physiological data acquisition was accomplished using a computer running eego™ software and using an eego amplifier (ANT Neuro, Enschede, Netherlands). The electrocardiogram (ECG) was recorded from three Ag/AgCl electrodes that were positioned on the participant’s chest in a modified lead II configuration. The ECG was recorded continuously for three minutes while participants were seated comfortably during undisturbed resting conditions and asked to fix their gaze to a cross presented at the center of the screen. Impedance was kept below 5 kΩ and each ECG signal was amplified, band-pass filtered (0.3–100 Hz) and sampled at 1000 Hz.

### 2.4. HRV analysis

In order to assess time- and frequency-domain HRV parameters, the ECG signal was analyzed offline using the Biopac Acqknowledge 5.0 software (Biopac Systems Inc., USA). All ECG data were visually inspected, and artefacts were removed. A digital trigger detecting R-waves was applied to the ECG signal to obtain RR intervals, corresponding to the inverse of heart rate. One participant in the group with dysphoria was excluded due to extended artefacts in the ECG signal. Then, time- and frequency-domain indices of HRV were calculated using Kubios HRV Analysis Software 3.3.1 (Matlab, Kuopio, Finland). Time-domain indices were calculated as follows:

1. Standard deviation of NN intervals (SDNN) expressed in ms, which reflects the cyclic components responsible for the variability of heart rate (Task Force, 1996);
2. Root mean square successive difference of NN intervals (rMSSD) expressed in ms, which reflects estimates of short-term variability of heart rate. rMSSD is highly sensitive to the fluctuation of high frequency of HRV and is an index of vagal control on heart (Task Force, 1996);
3. Number of interval differences of successive NN intervals greater than 50 ms in the entire recording (NN50), which reflects estimates of short-term variability of heart rate and is an index of vagal control on heart (Task Force, 1996).

Among time-domain HRV indices, SDNN, rMSSD, and NN50 were calculated as the most appropriate HRV measures for short-term recordings (Task Force, 1996) and as the most frequently used measures in literature (e.g., Kleiger et al., 1987; Koch et al., 2019; Patron et al., 2012; Patron et al., 2014).

Frequency-domain indices were obtained through autoregressive (AR) spectral analysis. AR method is a popular tool for spectral analysis of HRV and the length of data required for analysis is shorter than fast Fourier transform. The frequency-domain indices were calculated as follows:

1. Very low frequency (VLF) power (0 to 0.04 Hz) in ms^2^, which reflects sympathetic and parasympathetic inputs on the heart and may be influenced by the renin angiotensin, thermoregulatory, and peripheral vasomotor systems (e.g., Kamath et al., 1987; Taylor et al., 1998);
2. Low frequency (LF) power (0.04 to 0.15 Hz) in ms^2^, which reflects both sympathetic and parasympathetic cardiac activity and is strongly related to blood pressure regulation (Kamath et al., 1987);
3. High frequency (HF) power (0.15 to 0.40 Hz) in ms^2^, which primarily reflects cardiac parasympathetic tone (Kamath et al., 1987; Task Force, 1996).

All time- and frequency-domain indices were logarithmically transformed to normalize their distribution. In addition, the LF/HF ratio was computed as the ratio of LF(ms^2^)/HF(ms^2^) as it is thought to be a measure of sympathovagal balance (Malliani et al., 1991; Task Force, 1996).

### 2.5. Statistical analysis

Chi-square analysis was conducted to compare the three groups in terms of gender and separate analyses of variance (ANOVAs) with Group (dysphoria, PDS, controls) as the between-subjects factor were conducted on age, years of education and smoking habits. Because the effect of gender was marginally significant (*X*^2^(2, 67) = 5.3, *p* = .07) and male population was slightly underrepresented in the group with dysphoria and with past depressive symptoms, gender was used as a covariate in the statistical analysis conducted in the present study. Therefore, separate analyses of covariance (ANCOVAs) with Group (dysphoria, PDS, controls) as a between-subjects factor and gender as a covariate were conducted on questionnaires scores and on time-domain (lnSDNN, lnrMSSD and lnNN50) and frequency-domain (lnVLF, lnLF, lnHF and LF/HF) HRV measures. The Kolmogorov–Smirnov test was utilized to assess the normality of the distribution. Significant main effects (*p* < .05) were followed by Tukey HSD post-hoc tests in order to correct for multiple comparisons. Effect size was reported with Cohen’s d (small: d = 0.2; medium: d = 0.5; large: d = 0.8; Cohen, 1988).

## 3. Results

### 3.1. Demographics and clinical characteristics

With respect to demographic variables, Chi-square analysis revealed no group difference for gender, χ^2^ = 5.3, *p* = .071, and the Fisher’s exact test revealed no group difference for age, F_(2,64)_ = 0.38, *p* = .687, *η*^*2*^ = .01, education, F_(2,64)_ = 0.43, *p* = 0.649, *η*^*2*^_*p*_ = .01 and smoking habits, F_(2,64)_ = 1.89, *p* = .159, *η*^*2*^_*p*_ = .06^1^. Regarding the psychological measures, a significant main effect of Group on BDI-II score, F_(2,63)_ = 63.6, *p* = < .001, *η*^*2*^_*p*_ = .67, was found. The group with dysphoria showed significantly higher BDI-II scores than the control group (*p* < .001) and the PDS group (*p* <.001), whereas controls and PDS individuals did not differ from one another (*p* = .997). Separate ANCOVAs yielded a significant main effect of Group on BAI scores, F_(2,63)_ = 17.7, *p* < .001, *η*^*2*^_*p*_ = .36 and ERQ-suppression scores, F_(2,63)_ = 6.2, *p* = .004, *η*^*2*^_*p*_ = .16. Specifically, the group with dysphoria reported higher scores in the two questionnaires than both control and PDS groups, whereas the PDS group equaled the values of the control participants in all the measures. The ERQ-reappraisal scores did not differ among groups, F_(2,63)_ = 2.2, *p* = .120, *η*^*2*^_*p*_ = .07. The sample’s demographic and psychological measures, along with specific statistics, are reported in Table 1.

**Table 1.** Demographic and psychological characteristics of the control, the past depressive symptoms (PDS) and the dysphoria groups.

| Variable | Control (*n* = 25) | PDS (*n* = 16) | Dysphoria (*n* = 26) | *p* |
| --- | --- | --- | --- | --- |
| **Age (years)** | 20.4 (1.7) | 19.8 (6.2) | 20.7 (2.5) | .687 |
| **Male gender n (%)** | 12 (48) | 4 (25) | 6 (23) | .071 |
| **Education (years)** | 14.9 (1.3) | 14.6 (1.1) | 14.9 (1.5) | .645 |
| **Smoking (cigarettes /per day)** | 0.7 (1.5) | 1.3 (2.7) | 1.9 (2.4) | 0.159 |
| **BDI-II** | 4.2 (2.5) | 4.1 (2.6) | 20.5 (8.4) | <.001<sup>b,c</sup> |
| **BAI** | 6.3 (5.5) | 6.5 (4.0) | 18.8 (10.9) | <.001<sup>b,c</sup> |
| **ERQ-Reappraisal** | 31.0 (3.8) | 26.8 (5.4) | 28.4 (7.8) | .120 |
| **ERQ-Suppression** | 12.0 (4.0) | 10.9 (5.1) | 15.8 (6.0) | .004<sup>b,c</sup> |

Notes: Data are reported *M* (*SD*) of continuous and *n* (%) of categorical variables.
<sup>a</sup> = significant difference between control and PDS
<sup>b</sup> = significant difference between control and dysphoria
<sup>c</sup> = significant difference between PDS and dysphoria

### 3.2. HRV measures

The Kolmogorov–Smirnov test was not significant (*ps* > .05) indicating that data were normally distributed. Separate ANCOVAs on HRV time-domain indices yielded a significant main effect of Group for the lnSDNN, F_(2,63)_ = 6.20, *p* = .003, *η*^*2*^_*p*_ = .16, and lnrMSSD, F_(2,63)_ = 3.37, *p* = .041, *η*^*2*^_*p*_ = .10. Specifically, Tukey HSD post-hoc test revealed that the group with PDS and the group with dysphoria showed significantly lower lnSDNN values than the control group (PDS vs. control, *p* = .018, Cohen’s d = 0.82; dysphoria vs. control, *p* = .006, Cohen’s d = 0.94), whereas no difference between the PDS and individuals with dysphoria emerged (*p* = .999), as shown in Figure 1 (panel b). In contrast, Tukey HSD post-hoc test revealed that lnrMSSD values did not differ between the group with dysphoria and the group with PDS (*p =* .950), whereas the control group showed marginally significant higher values than the group with dysphoria (*p* = .080, Cohen’s d = 0.63) and the group with PDS (*p* = .070, Cohen’s d = 0.71). No significant main effect of Group on lnNN50 was noted (*p* = .087).

**Figure 1.**
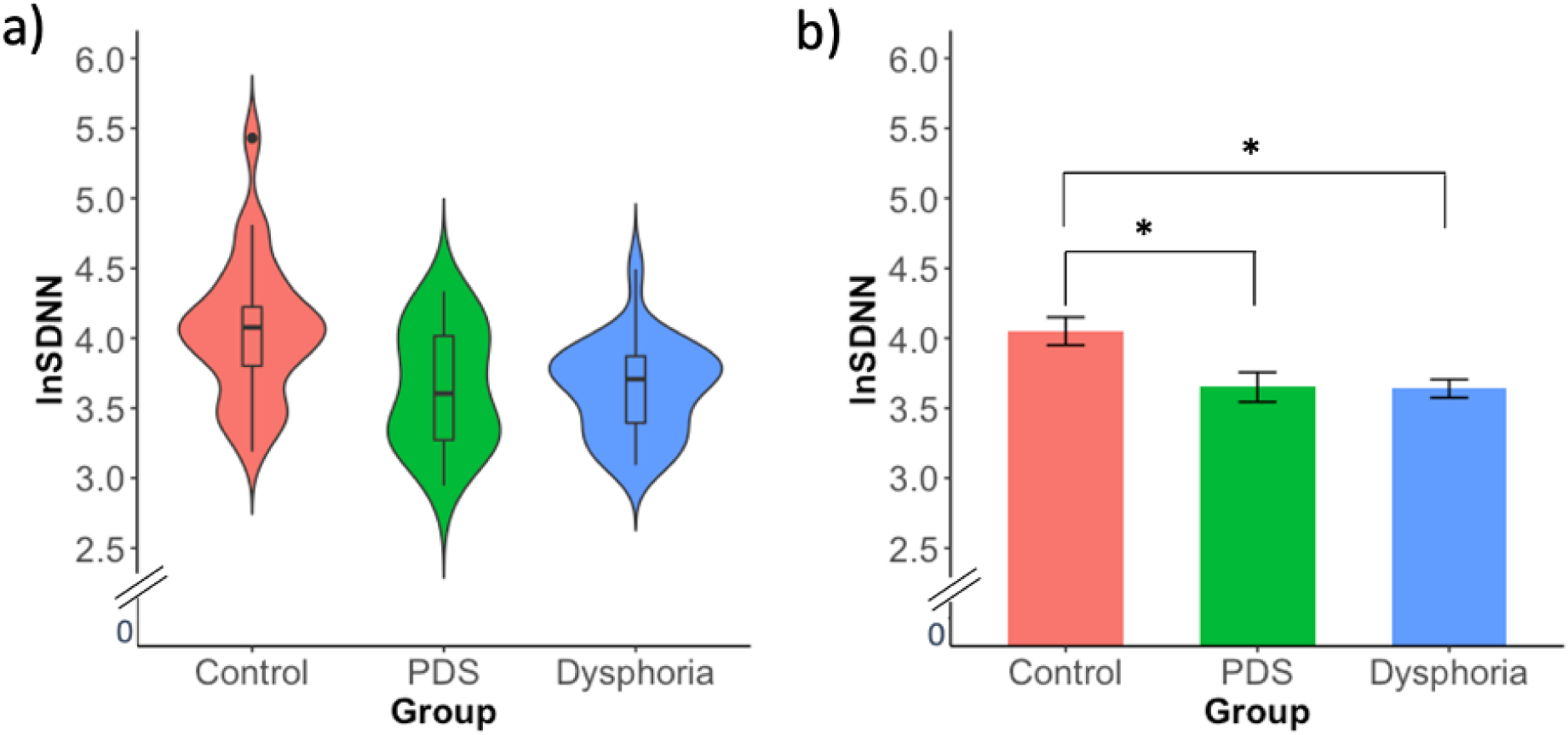
**Panel a)** Violin plot of the distribution of lnSDNN values for the three groups (control, PDS and dysphoria). The plot shows the median (indicated by the black horizontal band), the first through the third interquartile range (the vertical band), and estimator of the density (thin vertical curves) of the lnSDNN in each group (comparable to a boxplot, except that the distribution of the variable is illustrated as density curves). The violin plot outlines illustrate kernel probability density, i.e. the width of the shaded area represents the proportion of the data located there. **Panel b)** Mean lnSDNN values for the three groups (control, PDS and dysphoria). Error bars represent ± standard error of the mean (SEM). *p < .05 Notes: PDS= past depressive symptoms

With respect to frequency-domain indices, separate ANCOVAs revealed a significant main effect of Group on lnHF power, F_(2,63)_ = 3.97, *p* = .024, *η*^*2*^_*p*_ = .11, and lnLF power, F_(2,63)_ = 3.91, *p* =.025, *η*^*2*^ _*p*_*>* = .11. Tukey HSD post-hoc revealed that both in the group with dysphoria and in the group with PDS lnHF values were lower than those of the control group (PDS vs. control, *p* = .046, Cohen’s d = 0.72; dysphoria vs. control, *p* = .049, Cohen’s d = 0.71), as shown in Figure 2 (panel b). Specifically, comparable lnHF values were observed in the PDS group and in the group with dysphoria (*p* = .954). Regarding lnLF values, the group with dysphoria showed significantly lower values than the control group (*p* = .020, Cohen’s d = 0.81), whereas no difference emerged between the group with dysphoria and the PDS group (*p* = .675). Moreover, PDS and control groups did not differ from each other (*p* = .234). No significant difference among groups on lnVLF power (*p* = .644) and on LF/HF ratio (*p* = .748) was noted. All raw scores of time- and frequency-domain HRV parameters are reported in Table 2. The distributions of lnSDNN and lnHF values, as well as the median, the first and third interquartile range and the estimator of density, are displayed in Figure 1 (panel a) and in Figure 2 (panel a), respectively.

**Table 2.** Time- and frequency-domain HRV measures in control, past depressive symptoms (PDS) and dysphoria groups.

| Variable | Control (*n* = 25) | PDS (*n* = 16) | Dysphoria (*n* = 26) |
| --- | --- | --- | --- |
| **Time-domain indices** |  |  |  |
| **SDNN (ms)** | 65.7 (41.8) | 41.7 (17.7) | 40.0 (14.2) |
| **rMSSD (ms)** | 62.0 (36.0) | 40.0 (20.0) | 42.1 (24.1) |
| **NN50 (count)** | 72.0 (42.9) | 43.5 (40.4) | 42.9 (21.4) |
| **Frequency-domain indices** |  |  |  |
| **VLF (ms<sup>2</sup>)** | 1021.0 (4571.3) | 83.5 (63.9) | 75.3 (49.4) |
| **LF (ms<sup>2</sup>)** | 1426.0 (1306.6) | 843.0 (744.2) | 595.0 (356.9) |
| **HF (ms<sup>2</sup>)** | 2268.0 (2667.0) | 945.0 (993.2) | 817.0 (546.1) |
| **LF/HF (ms<sup>2</sup>)** | 1.7 (2.3) | 1.6 (1.6) | 1.1 (1.0) |

Notes: Results are reported as *M* (*SD*). HRV = heart rate variability; SDNN = standard deviation of all NN intervals in ms; rMSSD = square root of the mean of the sum of the squares of differences between adjacent NN intervals in ms; NN50 = number of pairs of adjacent NN intervals differing by more than 50 ms in the entire recording; VLF = very low frequency power; LF = low-frequency power; HF = high-frequency power; LF/HF = the ratio LF(ms<sup>2</sup>)/HF(ms<sup>2</sup>). *Note.* Data are *M* (*SD*).

**Figure 2.**
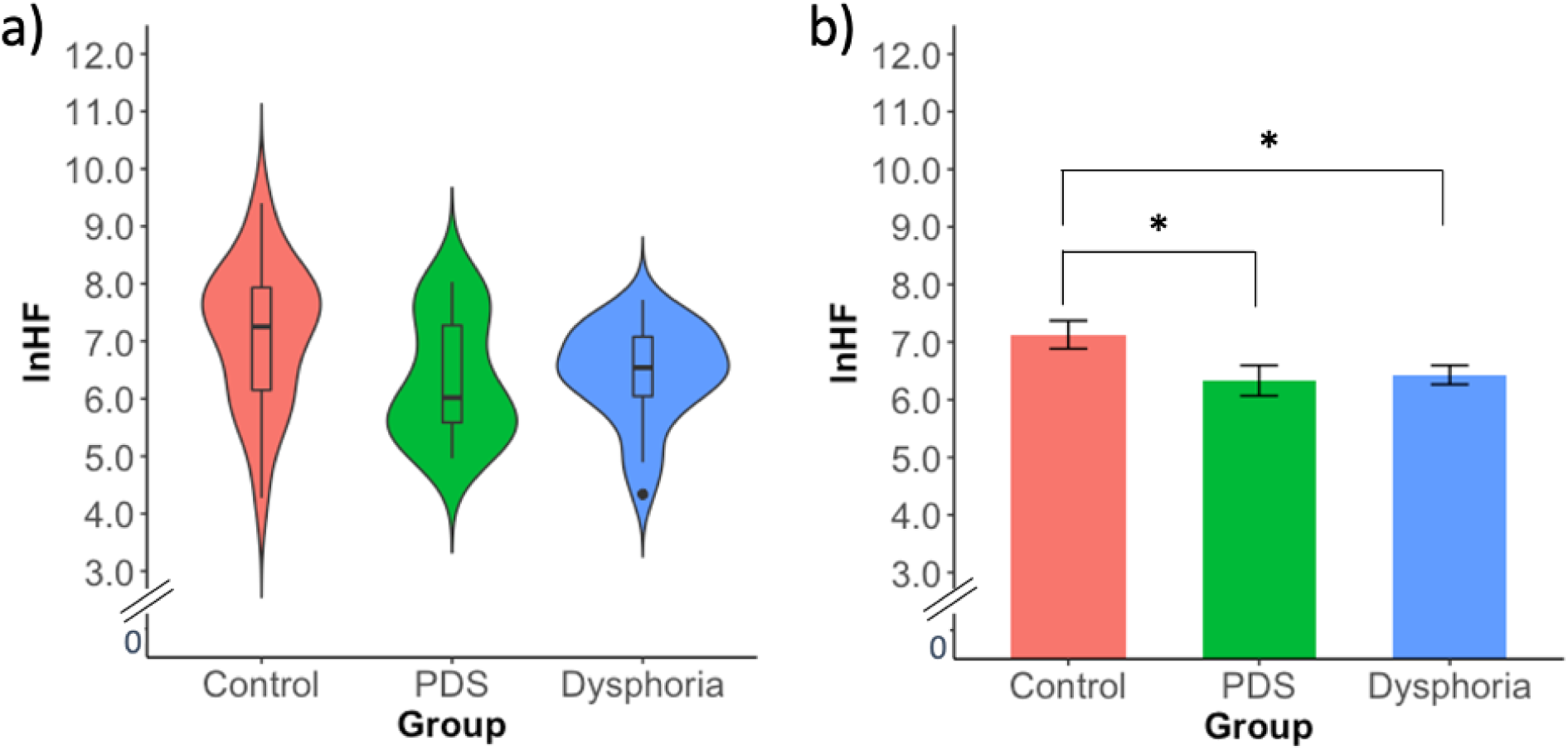
**Panel a)** Violin plot of the distribution of lnHF values for the three groups (control, PDS and dysphoria). The plot shows the median (indicated by the black horizontal band), the first through the third interquartile range (the vertical band), and estimator of the density (thin vertical curves) of the lnHF in each group (comparable to a boxplot, except that the distribution of the variable is illustrated as density curves). The violin plot outlines illustrate kernel probability density, i.e. the width of the shaded area represents the proportion of the data located there. **Panel b)** Mean lnHF values for the three groups (control, PDS and dysphoria). Error bars represent ± standard error of the mean (SEM). *p < .05. Notes: PDS= past depressive symptoms

## 4. Discussion

The present study aimed to investigate whether reduced HRV could be a potential indicator of vulnerability to depression. The intention was to contribute to the literature that aims at shedding light on biological factors involved in the development and maintenance of mood disorders. Particularly, the ANS is fundamental to wellbeing and is predominantly modulated by mechanisms of the central nervous system associated with several psychological functions, such as emotion regulation and general mental flexibility (Thayer and Lane, 2009). Understanding whether ANS modifications are present in subclinical and post-acute phases of depression is important for early recognition and prevention of depression (Basset et al., 2016). To this end, resting state HRV was analyzed in two groups (with dysphoria and with past depressive symptoms) known to be both at-risk conditions of developing a full-blown depressive episode, in comparison to a group of control participants. This was done because, although many studies documented reduced HRV during acute depression (Brunoni et al., 2013; Hartmann et al., 2019; Jangpangi et al., 2016; Licht et al., 2008; Nahshoni et al., 2004; Udupa et al., 2007; for a review see Kemp et al., 2010), studies addressing at-risk conditions were lacking.

In line with our hypothesis, individuals with dysphoria and those with past depression showed reduced cardiac vagal modulation compared to control participants. For instance, results showed significant reductions in both SDNN and HF, with medium-to-large effect sizes. In addition, the two groups also showed marginally significant rMSSD, compared to control participants. Specifically, there seems to be a trend in rMSSD estimates that is similar to those of the HF, confirmed by a medium-to-large effect size. Although rMSSD is typically correlated with the HF power, rMSSD is partly influenced by lower frequency fluctuations (< 0.12 Hz; Berntson et al., 2005), outside the HF power range, which may explain the slightly different result between rMSSD and HF power. In regard to LF, individuals with dysphoria showed reduced values compared to both control and individuals with past depression. Contrarily, LF/HF ratio did not differ between the three groups.

It is worth noting that individuals with dysphoria and past depression did not differ, showing comparable HF and SDNN parameters. These findings suggest that the two at-risk groups were characterized by reduced vagally mediated cardiac modulation rather than increased sympathetic cardiac modulation. Indeed, considering that resting heart rate variability is mostly coordinated by the parasympathetic system, the SDNN, which reflects the cyclic components responsible for the variability of heart rate (Task Force, 1996), is more likely to reflect cardiac vagal control rather than sympathetic control (Zaza and Lombardi, 2001). Likewise, extensive experimental and clinical evidence maintains that HF power is mainly supported by vagal activity. On the other hand, the role of VLF, which did not differ between groups, is controversial and cannot be clearly attributed to a specific autonomic modulation (Zaza and Lombardi, 2001). Altogether, the current findings suggest that vulnerability to depression is associated with reduced vagal rather than increased sympathetic modulation of the heart at rest. Vagal control of the heart is crucial not only in the regulation of physiological functions (i.e. immune, inflammatory, and cardiac functions; Thayer and Sternberg, 2006; Weber et al., 2010), but also in the communication between the brain’s integrative system for adaptive regulation and the periphery (Hansen et al., 2004; Thayer et al., 2012). Correspondingly, reduced parasympathetic activity has been associated with less emotion regulation skills (Thayer and Lane, 2009) and reduced mental flexibility (Carnevali et al., 2018), which, in turn, have been associated with higher risk of depression.

In regard to the group with past depressive symptoms, although the literature is fairly limited, our results are in line with previous findings that reported reduced HRV in individuals with past depression (Bär et al., 2004; Basset et al., 2016; Brunoni et al., 2013; Licht et al., 2008). However, some investigations on reduced HRV in individuals with past depression have also reported null findings (Ahrens et al., 2008), or findings limited to a subgroup with a history of suicidal ideation (Chang et al., 2013). It has to be noted that available studies on this matter differ in samples (i.e., participants age) and applied methodology (i.e., HRV domains). Importantly, most of the previous studies included individuals who had taken antidepressant medication in the past, which may be a confounding factor and explain at least part of the inconsistencies in HRV findings. In contrast, in the present study, both at-risk samples were free from any current and past medications, thus removing this potential confounding factor.

With respect to the group with dysphoria, the literature is extremely limited. Yet, our results are in line with previous findings showing reduced parasympathetic activity in individuals with somatic symptoms of depression (Bosch et al., 2009; Messerotti Benvenuti et al., 2015) and in adolescents with subclinical depressive symptoms (Blood et al., 2015). Additionally, a longitudinal study on a non-clinical sample reported that the presence of depressive symptoms was correlated with reduced HRV on a time frame of almost three years, suggesting that low vagal tone may be prospectively implicated in the generation of depressive symptoms (Carnevali et al., 2018). Furthermore, a recent prospective study showed that healthy men with increased HRV were less likely to develop depressive symptoms at 10 years follow-up (Jandackova et al., 2016).

While HRV was significantly reduced in the group with past depression compared to controls, the two groups did not show any significant change in the investigated psychological measures. On the other hand, the group with dysphoria presented alterations both in HRV and in all the investigated psychological measures. These findings are in line with the extensive literature that described reduced HRV as a transdiagnostic correlate of psychopathology. As a matter of fact, reduced HRV was also observed in bipolar disorder and post-traumatic stress disorder (Carr et al., 2018; Smith et al., 2020). A reason for this is that HRV reflects, albeit peripherally, prefrontal cortex functioning and, therefore, captures the psychophysiological vulnerability that many psychopathologies share (Beauchaine and Thayer, 2015). Besides, the present study not only is in line with this conceptualization of HRV, but it also showed preliminary evidence that HRV could be a potential vulnerability factor to depression. In line with this, future studies should focus on determining whether reduced HRV could be a vulnerability factor to other psychopathologies beyond depression.

Moreover, the evidence that HRV might embody a vulnerability factor for the development of psychopathology raises the issue of whether in addition to be a trait-dependent feature, it may also be a state-dependent feature, associated with situational demands. Indeed, a series of studies have demonstrated that, not only baseline, but even task-related HRV could represent an index of clinical outcome in individuals with depression (Shinba et al., 2014; Shinba et al., 2020). Although resting-state HRV is a well-validated and feasible tool for the clinical practice, state-dependent HRV-fluctuations may increase the accuracy of HRV as a candidate endophenotype of depression.

From a clinical perspective, the current data suggest that reduced cardiac vagal modulation could represent a potential indicator of vulnerability to depression. Hence, HRV may serve as an indicator of risk of clinical depression, providing a valuable tool for the early recognition or post-depressive monitoring of vulnerable individuals. This will increase clinicians’ ability to identify individuals at risk for depression who can benefit from targeted and indicated prevention by psychiatric and psychological interventions. Conventional treatments for depression (i.e., antidepressant and psychotherapy), that usually improve psychological wellbeing (Kemp et al., 2010; Licht et al., 2010), do not seem to improve cardiac vagal tone. Inversely, it has been shown that tricyclic antidepressants could lead to a further reduction of HRV, in addition to be effective in only a third of a half of patients (Garcia-Toro et al., 2012; Hughes and Cohen, 2009). In this context, considering that standard treatment does not guarantee normalization of the ANS activity (Kemp et al., 2010), the demand to consider alternative evidence-based interventions to improve cardiac autonomic modulation is increasingly recognized. A widely applied intervention used to increase cardiac vagal control is HRV-biofeedback, a non-invasive technique that allows psychophysiological self-regulation. HRV-biofeedback works by providing a direct feedback of HRV (through a screen or headphones), which helps individuals to actively modify that targeted physiological process (Lehrer, 2007). HRV-biofeedback has been found to be effective in restoring cardiac autonomic balance, by reducing sympathetic over-reactivity and increasing cardiac vagal tone, that, in turn, improves wellbeing by reducing stress responses (Lehrer, 2007). Accordingly, promising evidence showed that HRV-biofeedback could be a successful adjunctive treatment for depression (Caldwell and Steffen, 2018; Karavidas et al., 2007; Patron et al., 2013; Siepmann et al., 2008; Zucker et al., 2009).

The current study presents strengths which distinguish it from other work in this area. First, individuals with neurological and cardiological conditions were excluded, as well as those under medications, thus avoiding potential confounding effects on HRV. Second, the comparative study on two at-risk groups allowed to examine the role of HRV as a potential vulnerability factor to depression adopting a comprehensive approach.

## 5. Limitations

In interpreting our results, several limitations have to be acknowledged. First, the inclusion of a group with clinical depression would have provided a complete overview about HRV during the illness course. Indeed, the inclusion of a group with clinical depression would have contributed to establishing whether HRV is an index capable of discriminating among different degrees of depression severity. Second, past depression was assessed retrospectively on the basis of self-reported information, without granting further data (i.e., past psychological evaluations, hospital records, family members’ reports). In addition, considering that one past episode is sufficient to categorize past depression, the number of episodes were not collected. Likewise, although all the participants did not report any medical condition and diagnosis during the anamnestic interview, it cannot be ruled out the presence of any psychiatric conditions. Indeed, the use of only the module A of the SCID-5-CV did not allow to investigate whether any participant would fit in with any psychiatric diagnoses beyond depressive or bipolar disorders. Also, the relatively small sample size, as well as the young age of the participants, might not allow generalization of the findings. However, the relatively small sample size was sufficient to achieve adequate statistical power (1-β = 0.88), demonstrating that the two at-risk groups were characterized by reduced vagally-mediated HRV by medium-to-large effect sizes. Also, the young age of the participants, which corresponds to the life phase between late adolescence and early adulthood, was selected because it is a known to be sensitive period for the development of depressive symptoms (Grano et al., 2015). Indeed, it is acknowledged to be a stage characterized by individual adjustments and life transitions, known to challenge individuals with higher rates of depression (Grano et al., 2015).

## 6. Conclusions

Taken all together, in the present study both at-risk samples (participants with dysphoria and past depression) presented with reduced vagal tone, indexed by lower HRV than healthy controls. In turn, these findings suggest that reduced HRV is likely to represent a potential endophenotype, implicated in the risk of developing full-blown depression, rather than being a mere correlate of current depressive state. A longitudinal evaluation examining the development of depressive symptoms and HRV is crucial to determine whether reduced HRV may predict the onset and recurrence of depression.

## Data Availability

data will be made available to researchers upon reasonable request.

## Footnotes

1 ^1^ Applying the classification daily smokers (at least one cigarette/per day) vs. non-smokers there was no significant difference between the three groups with Chi-square analysis (*p* = 0.18)

